# Racial and Ethnic Differences in Exposure to Antibiotics Associated with *Clostridioides difficile* Infection in US Academic Dental Care

**DOI:** 10.64898/2026.06.25.26356622

**Authors:** Adrianne D. Gladden, Leo K. Westgard, Rachel A. Tam, Martin C. Ugbala, Kap Sum Foong, Alysse G. Wurcel

**Affiliations:** Department of Molecular Biology and Microbiology, Tufts University, Boston, Massachusetts, 02111, USA; Graduate School of Biomedical Sciences, Tufts University, Boston, Massachusetts, 02111, USA; Independent Research Consultant (affiliated with Boston Medical Center), Boston, MA, USA; Department of Medicine, Section of General Internal Medicine, Boston Medical Center, Boston, MA, USA; UTHealth Houston School of Dentistry, Houston, Texas, USA; Department of Medicine, Division of Infectious Diseases, Denver Health, Denver, CO, USA; Boston University Chobanian & Avedisian School of Medicine, Boston, MA, USA

## Abstract

**Background:** Severe *Clostridioides difficile* infection (CDI) morbidity and mortality disproportionately affect Black and Hispanic patients in the United States. Antibiotic exposure is the primary modifiable risk factor for CDI, and clindamycin is among the agents most strongly associated with related harm. Characterizing inequities in prescribing is critical. Dentistry is a major source of clindamycin prescriptions. Academic dental clinics serve diverse patient populations and provide an ideal setting to evaluate prescribing across racial and ethnic groups. We therefore examined antibiotic use and cumulative clindamycin exposure as measures of CDI-associated risk.

**Methods:** We conducted a retrospective study of electronic health records from 5 US academic dental institutions from 2021 through 2023. We analyzed 552,428 encounters among 132,770 patients with documented race/ethnicity to estimate adjusted odds of receiving any oral antibiotic and clindamycin by race/ethnicity. Secondary outcomes evaluated total antibiotic exposure among dental provider–prescribed antibiotics, focusing on higher-than-standard cumulative dosing of clindamycin (>8400 mg) and amoxicillin (>10,500 mg).

**Results:** Oral antibiotic prescribing occurred in 1.9% of encounters. Compared with White patients, Black, Hispanic, and Other race patients had slightly lower adjusted odds of receiving any oral antibiotic, while Black patients had greater odds of receiving a higher-than-standard cumulative clindamycin dose when clindamycin was prescribed (adjusted odds ratio, 2.19; 95% confidence interval, 1.25–3.82).

**Conclusion:** Racial and ethnic inequities in dental antibiotic prescribing extended beyond antibiotic receipt to cumulative clindamycin exposure. Although CDI outcomes were not directly measured, these prescribing differences may have implications for disparities in CDI-associated harm and warrant further investigation.

## BACKGROUND

Severe *Clostridioides difficile* infection (CDI) disproportionately burdens Black and Hispanic patients in the United States with higher rates of complications and prolonged hospitalization, whereas White populations historically exhibit higher documented population-level incidence.^1–3^ Prior outpatient antibiotic exposure precedes approximately 60-65% of community-associated (CA) CDI cases and is a major predictor of both CDI incidence and clinical severity.^4^

Clindamycin is consistently among the antibiotics associated with the highest risk of CDI and severe CDI outcomes.^5,6^ Although even a single clindamycin dose may precipitate clinically significant disease, higher cumulative exposure to antibiotics confers greater risk.^7–10^ Despite this, antibiotic stewardship efforts often focus primarily on antibiotic selection, such as discouraging clindamycin use, while incompletely accounting for cumulative exposure through dose and duration.^11,12^

Dentistry represents a major source of outpatient clindamycin exposure in the United States (US) and has been linked to a substantial proportion of antibiotic-associated CA-CDI cases.^13–17^ Given the association between cumulative clindamycin exposure and CDI risk, differential upstream prescribing patterns may contribute to downstream racial and ethnic disparities in CDI vulnerability and severity.

Disparities in outpatient antibiotic prescribing are well documented, including in dentistry.^18,19^ Prior studies suggest that Black and Hispanic patients are more likely to receive guideline-discordant antibiotics, and that inappropriate dental antibiotic prescribing frequently involves clindamycin.^18,20^ Disproportionate clindamycin prescribing to Black patients has also been observed in at least one clinical setting.^21^ However, less is known about whether racial and ethnic differences exist in cumulative clindamycin exposure that may have important implications for CDI vulnerability beyond antibiotic receipt alone. To investigate this possibility, we analyzed antibiotic prescribing patterns across academic dental institutions spanning multiple US geographic regions.

## METHODS

### Study Design and Cohort Definition

We conducted a retrospective study using deidentified electronic health record data from 5 US academic dental institutions contributing to the BigMouth Dental Data Repository. BigMouth Dental Data Repository is a centralized, multi-institutional database containing partially deidentified electronic health record data from academic dental institutions across the US and has been used in prior studies of dental care and prescribing patterns.^22,23^ The analytic period spanned January 1, 2021 through December 31, 2023.

Analyses were performed at the encounter level. An encounter was defined as all dental visits for a given patient within a single calendar month, reflecting the level of temporal granularity available in the dataset. Primary analyses were restricted to records with documented race/ethnicity, age, and sex.

We harmonized race and ethnicity across institutions using available structured fields. Hispanic or Latino ethnicity was prioritized and was treated as mutually exclusive; all remaining designations reflected non-Hispanic individuals. Patients were assigned to White, Black, Hispanic, Asian, Other, or Unknown. The Other category included American Indian or Alaska Native, Native Hawaiian or Other Pacific Islander, Multiracial, and Some Other Race. Records with Unknown race/ethnicity were retained for descriptive analyses but excluded from primary adjusted regression analyses.

Patient-level characteristics included age, sex, treating institution, documented antibiotic allergy, and selected comorbidities (diabetes and chronic obstructive pulmonary disease). ZIP3 was defined as the 3-digit prefix of the patient ZIP code. For each site, the majority ZIP3 was used to determine its US geographic region.

### Antibiotic Classification and Dose Computation

Prescription records included heterogeneous medication entries derived from structured and free-text fields. We standardized medication names by mapping generic, branded, and formulation-specific terms to common antibiotic categories relevant to outpatient dental care, including amoxicillin, clindamycin, azithromycin, cephalexin, penicillin, metronidazole, doxycycline, and selected less commonly prescribed agents. Amoxicillin-clavulanate (Augmentin) was grouped with amoxicillin for classification and dose summarization.

We categorized route of administration using medication names and structured prescription fields as oral, topical, other, or unknown. Primary analyses focused on oral antibiotic prescribing. Records without an antibiotic prescription (“N/A”) were classified as no antibiotic exposure.

When structured prescription fields were available, total prescribed antibiotic dose was estimated using strength and quantity dispensed. For solid oral formulations, total dose was calculated as unit strength multiplied by quantity dispensed. Antibiotic-specific quantity upper limits were applied to identify implausible values that were potentially erroneous entries (e.g., amoxicillin, 250 units; clindamycin, cephalexin, and penicillin, 120 units; doxycycline and metronidazole, 180 units). For liquid formulations, total dose was estimated from concentration (mg/mL) multiplied by dispensed volume when interpretable quantities up to 1000 mL were available. When structured fields were incomplete, prescription instructions were used to estimate missing dose or quantity information, including entries requiring back-calculation or fields that appeared to represent total milligrams. Total dose was calculated for 26,236 of 26,237 (99.996%) oral antibiotic prescriptions, including 100% of amoxicillin and clindamycin prescriptions.

Calculated total doses were retained only if greater than 0 and no more than 120,000 mg. Because structured dose fields were not uniformly available across institutions, dose-based analyses were restricted to prescriptions with sufficient information to estimate total dose. Self-reported medication entries were excluded from total dose calculations.

We prespecified higher-than-standard cumulative dose thresholds based on American Dental Association clinical practice guidelines for the management of odontogenic infections, which recommend clindamycin 300 mg 4 times daily and amoxicillin 500 mg 3 times daily for 3 to 7 days.^20^ These recommendations correspond to maximum cumulative doses of 8,400 mg for clindamycin and 10,500 mg for amoxicillin.

### Antibiotic Outcomes

Primary outcomes were (1) receipt of any oral antibiotic during an encounter and (2) receipt of clindamycin among encounters with an antibiotic prescription. Dose-based outcomes were evaluated among encounters involving oral amoxicillin or clindamycin prescriptions. Higher-than-standard cumulative exposure was prespecified as total amoxicillin dose >10,500 mg and total clindamycin dose >8,400 mg.

### Clinical Procedure Context

Current Dental Terminology (CDT) procedure codes were grouped into broad clinical categories based on code prefix. To prioritize the clinical context most relevant to antibiotic prescribing, encounters with multiple procedure categories were assigned a primary category according to the following descending hierarchy: oral and maxillofacial surgery, endodontic, periodontal, restorative, prosthodontic, implant/prosthodontic, preventive, diagnostic, orthodontics, adjunctive/other, and other/unknown.

### Statistical Analysis

Descriptive statistics summarized patient- and encounter-level characteristics. Continuous variables are presented as medians with interquartile ranges (IQR) and categorical variables as counts and percentages.

We used multivariable logistic regression to evaluate associations between race/ethnicity and oral antibiotic receipt, clindamycin selection, and higher-than-standard cumulative amoxicillin and clindamycin exposure. To account for potential correlation among multiple encounters contributed by the same patient, we estimated cluster-robust standard errors at the patient level. Results are presented as adjusted odds ratios (aORs) with 95% confidence intervals (CIs). Primary models were restricted to encounters with known race/ethnicity and adjusted for age, sex, institution, calendar year, and primary procedure category.

### Robustness and Sensitivity Analyses

We performed sensitivity analyses to evaluate whether the association between Black race and higher-than-standard cumulative clindamycin dose differed across institutions. Models were fit separately by site, and a pooled interaction model was used to assess heterogeneity by institution.

To examine the impact of Unknown race/ethnicity, we conducted a probability-based reassignment analysis. Encounters with Unknown race/ethnicity were reassigned across the observed race/ethnicity categories using a multinomial model based on age, sex, institution, calendar year, and primary procedure category. In successive scenarios, the probability of reassignment to the Black category was progressively increased relative to the remaining categories, after which probabilities were rescaled and the primary adjusted model was re-estimated.

We also compared encounters with known versus Unknown race/ethnicity descriptively within institutions, including site-specific event rates and crude odds ratios for higher-than-standard cumulative clindamycin dose.

Finally, we performed a secondary descriptive analysis to determine how often encounters with dentist-prescribed antibiotics also included documentation of additional antibiotic exposure elsewhere in the electronic health record during the same encounter, including clindamycin specifically, to assess whether cumulative antibiotic exposure extended beyond the indexed dental prescription.

All analyses were performed in R, version 4.5.1.

### Ethics Statement

The Tufts Health Sciences Institutional Review Board determined that this study did not involve human subjects research and that IRB approval was not required (IRB ID STUDY00004869).

## RESULTS

### Study Population

The analytic cohort comprised electronic health records for 552,428 encounters during 2021-2023 from 132,770 unique patients with documented race/ethnicity. The proportion of patients with Unknown race/ethnicity varied substantially across institutions, ranging from 25.4% to 51.4% (Table 1).

**Table 1.** Patient-level cohort characteristics by institution, 2021-2023.

| Region | Multi-region | Northeast | West | West | Midwest | South |
| --- | --- | --- | --- | --- | --- | --- |
| Patients, No. | 225,348 | 20,771 | 49,632 | 25,808 | 83,279 | 45,858 |
| **Demographic characteristics** |  |  |  |  |  |  |
| Age, median (IQR), y | 38.0 (19.0, 60.0) | 43.0 (29.0, 61.0) | 35.0 (11.0, 62.0) | 45.0 (28.0, 63.0) | 32.0 (17.0, 56.0) | 41.0 (22.0, 59.0) |
| **Sex** |  |  |  |  |  |  |
| Female | 53.9% | 56.6% | 52.0% | 52.7% | 53.5% | 56.3% |
| Male | 45.0% | 42.7% | 46.4% | 47.3% | 45.6% | 42.5% |
| Other | 0.1% | 0.1% | 0.0% | 0.1% | 0.3% | 0.0% |
| Unknown | 0.9% | 0.6% | 1.6% | 0.0% | 0.7% | 1.1% |
| **Race/ethnicity** |  |  |  |  |  |  |
| White | 31.3% | 26.7% | 20.4% | 36.4% | 46.0% | 15.3% |
| Black | 7.5% | 5.6% | 4.5% | 10.7% | 6.6% | 11.5% |
| Hispanic | 13.7% | 6.1% | 18.6% | 22.2% | 5.3% | 22.5% |
| Asian | 4.0% | 8.8% | 2.6% | 4.2% | 3.0% | 5.2% |
| Other | 2.4% | 4.9% | 2.4% | 1.1% | 1.7% | 3.2% |
| Unknown | 41.1% | 47.8% | 51.4% | 25.4% | 37.4% | 42.4% |
| **Allergy** |  |  |  |  |  |  |
| Antibiotic allergy | 5.9% | 8.6% | 6.3% | 0.3% | 7.5% | 4.4% |
| **Comorbidity** |  |  |  |  |  |  |
| Diabetes | 15.9% | 7.7% | 15.1% | 11.4% | 7.8% | 37.7% |
| COPD | 1.6% | 0.3% | 1.1% | 1.5% | 1.4% | 3.3% |

Cohort was restricted to patients with at least one encounter from 2021–2023. Age is presented as median (IQR); categorical variables are presented as column percentages. Antibiotic allergy and comorbidity rows indicate the proportion of patients with a documented 'Yes.' Race/ethnicity was harmonized from structured race and ethnicity fields and includes an explicit Unknown category. Of 225,348 patients in the descriptive cohort, 132,770 (58.9%) had documented race/ethnicity and were eligible for adjusted regression analyses.

Participating sites represented the Northeast, West, Midwest, and South based on patient ZIP3 catchment areas. Median age ranged from 32.0 to 45.0 years, and 52.4% to 56.8% of patients were female. Demographic, clinical, and ZIP3-linked socioeconomic characteristics varied across institutions and racial/ethnic groups (Table 1; Table S1).

### Primary Antibiotic Prescribing Outcomes

After adjustment for age, sex, institution, calendar year, and primary procedure category, racial and ethnic differences in antibiotic prescribing were observed (Figure 1; Table 2). The overall rate of oral antibiotic prescription was 1.9% (Table S2). Compared with White patient encounters, Black patient encounters had lower odds of receiving any oral antibiotic (aOR, 0.85; 95% CI, 0.79–0.91). No significant association was observed between Black race and clindamycin selection among antibiotic-prescribing encounters (aOR, 0.85; 95% CI, 0.67–1.06). Hispanic patient encounters had lower odds of receiving any oral antibiotic (aOR, 0.80; 95% CI, 0.75–0.84) and lower odds of receiving clindamycin (aOR, 0.55; 95% CI, 0.43–0.68). Adjusted associations for prescribing outcomes across all race/ethnicity categories are shown in Table 2; complete multivariable model results are provided in Table S3.

**Figure 1.**
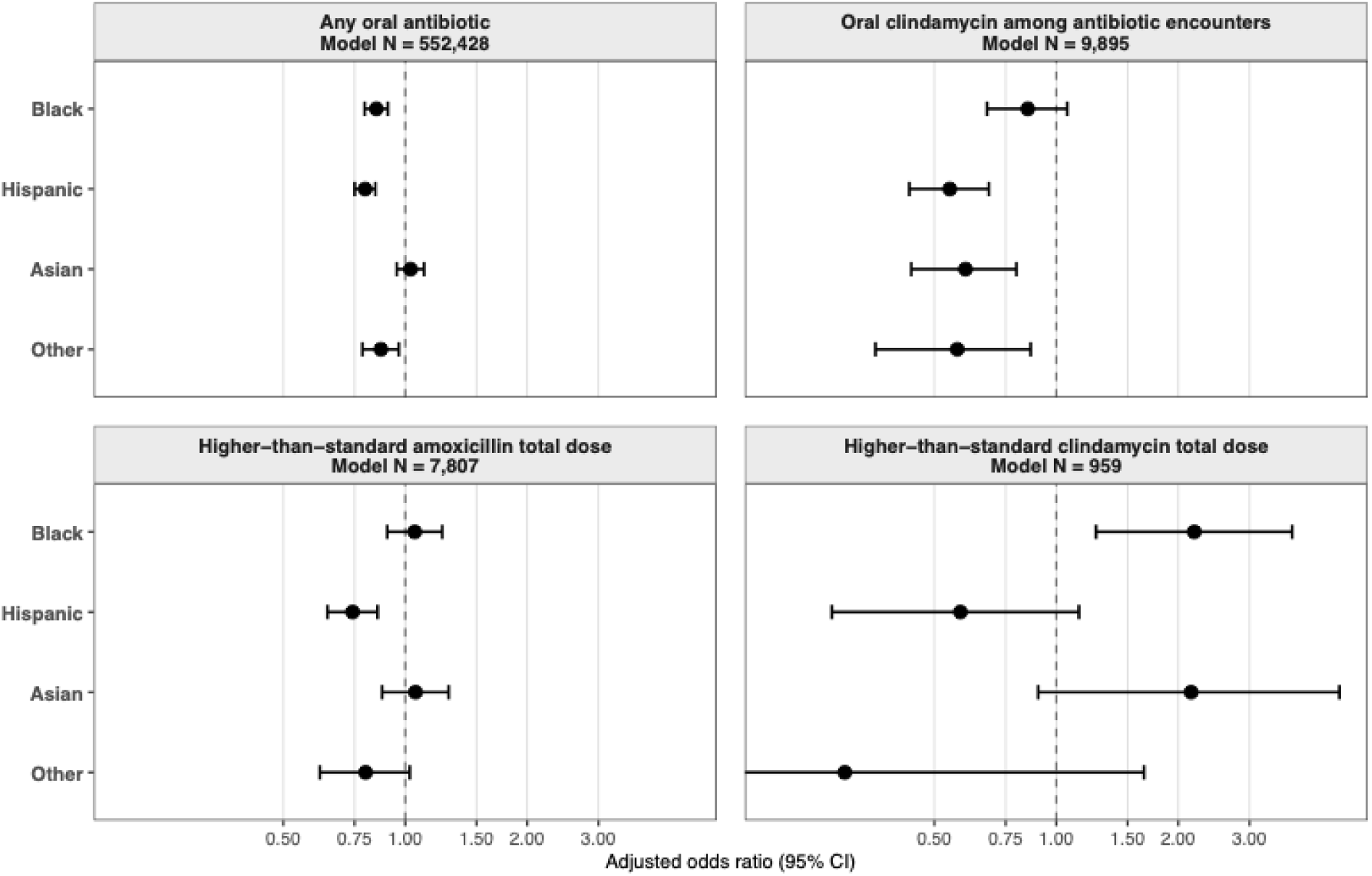
Adjusted associations of race/ethnicity with encounter-level antibiotic prescribing outcomes. Forest plots show adjusted odds ratios (ORs) and 95% confidence intervals (CIs) for encounter-level antibiotic prescribing outcomes, with White patient encounters as the reference group. Models were adjusted for age, sex, institution, calendar year, and primary procedure category. The oral clindamycin model was restricted to encounters with any oral antibiotic prescription. Higher-than-standard cumulative dose was defined as >10,500 mg for amoxicillin and >8,400 mg for clindamycin. Self-reported antibiotic entries were excluded from dose-based analyses. Analyses were restricted to encounters with known race/ethnicity.

**Table 2.** Adjusted associations of race/ethnicity with encounter-level antibiotic prescribing outcomes.

| Outcome | Model N | Race/ethnicity | Adjusted OR (95% CI) | P value |
| --- | --- | --- | --- | --- |
| Any oral antibiotic | 552,428 | Black | 0.85 (0.79–0.91) | <.001 |
|  |  | Hispanic | 0.80 (0.75–0.84) | <.001 |
|  |  | Asian | 1.03 (0.95–1.11) | .444 |
|  |  | Other | 0.87 (0.78–0.96) | .009 |
| Oral clindamycin among antibiotic encounters | 9,895 | Black | 0.85 (0.67–1.06) | .163 |
|  |  | Hispanic | 0.55 (0.43–0.68) | <.001 |
|  |  | Asian | 0.60 (0.44–0.80) | <.001 |
|  |  | Other | 0.57 (0.36–0.86) | .012 |
| Higher-than-standard amoxicillin total dose | 7,807 | Black | 1.06 (0.90–1.23) | .494 |
|  |  | Hispanic | 0.74 (0.64–0.85) | <.001 |
|  |  | Asian | 1.06 (0.88–1.28) | .541 |
|  |  | Other | 0.80 (0.62–1.03) | .084 |
| Higher-than-standard clindamycin total dose | 959 | Black | 2.19 (1.25–3.82) | .006 |
|  |  | Hispanic | 0.58 (0.28–1.14) | .126 |
|  |  | Asian | 2.15 (0.90–5.00) | .077 |
|  |  | Other | 0.30 (0.02–1.65) | .260 |

White patient encounters were the reference group. Models were adjusted for age, sex, institution, calendar year, and primary procedure category. The oral clindamycin model was restricted to oral antibiotic–receiving encounters. Higher-than-standard amoxicillin dose was defined as greater than 10,500 mg, and higher-than-standard clindamycin dose as greater than 8,400 mg. Self-reported antibiotic entries were excluded from dose-based analyses. Analyses were restricted to encounters with known race/ethnicity.

In dose-based analyses, higher-than-standard cumulative amoxicillin exposure occurred in 2,517 of 7,842 encounters (32.1%), whereas higher-than-standard cumulative clindamycin exposure (>8,400 mg) occurred in 138 of 962 encounters (14.3%) (Table S2). Compared with White patient encounters, Black patient encounters had higher adjusted odds of receiving higher-than-standard cumulative clindamycin doses (aOR, 2.19; 95% CI, 1.25-3.82). In contrast, Black patient encounters were not associated with higher-than-standard cumulative amoxicillin exposure after adjustment (aOR, 1.06; 95% CI, 0.90–1.23), although Hispanic patient encounters had lower adjusted odds relative to White patient encounters (aOR, 0.74; 95% CI, 0.64–0.85) (Table 2).

### Sensitivity Analysis

Frequencies of oral antibiotic prescribing, clindamycin selection, and higher-than-standard cumulative dosing varied across participating institutions (Table S2). We therefore evaluated site-stratified models to determine whether the higher odds of above-standard cumulative clindamycin dose observed in the pooled multicenter analysis were driven by a single institution. Two institutions had no above-standard clindamycin dosing events among Black patient encounters, limiting site-specific estimation. Among institutions where estimation was possible, site-specific odds ratios were consistently greater than 1. A pooled race-by-institution interaction model did not identify evidence of heterogeneity by institution (interaction P = .49) (Figure 2A).

**Figure 2.**
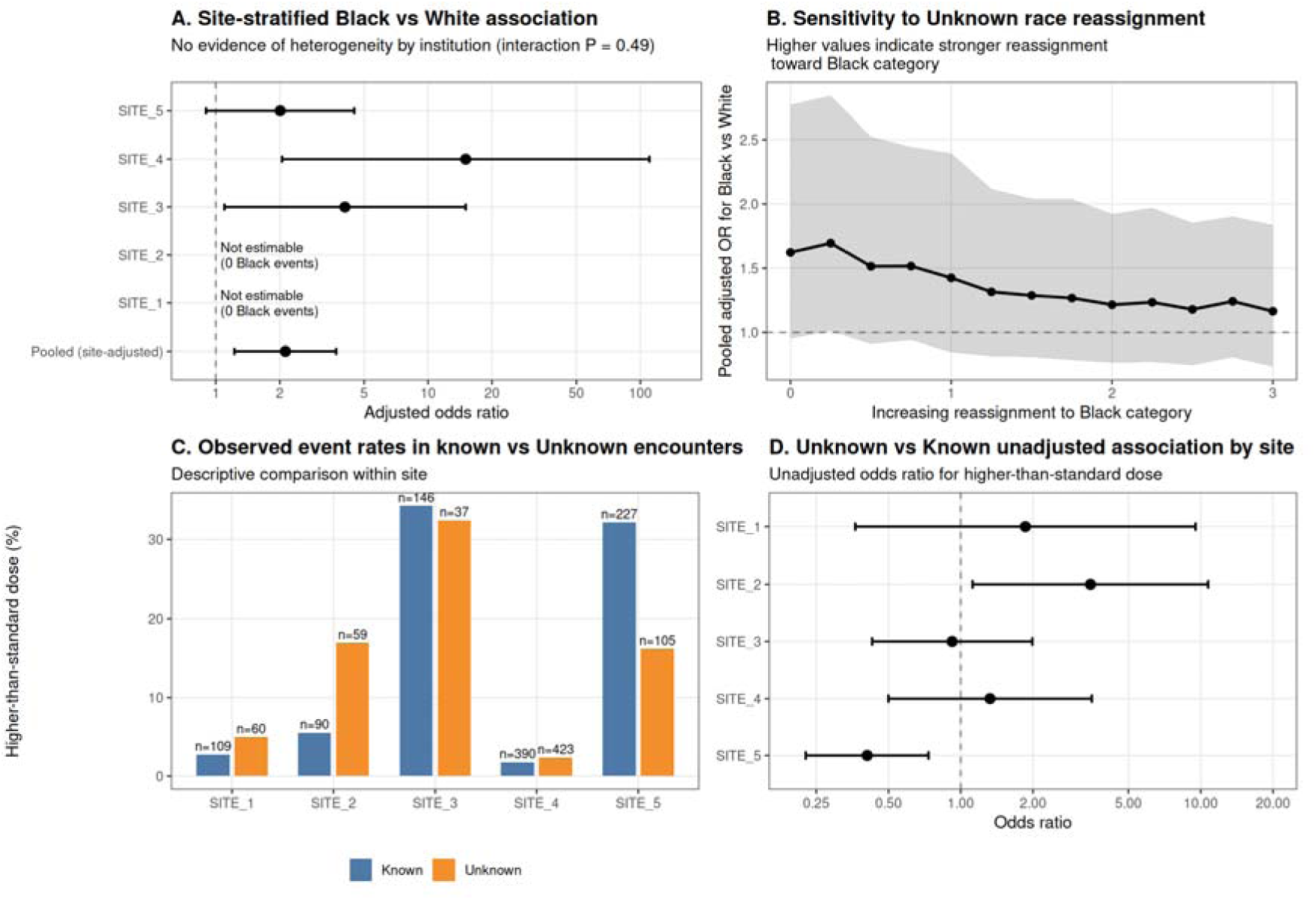
Robustness analyses for higher-than-standard cumulative clindamycin dose. Panel A shows pooled and site-specific adjusted odds ratios (ORs) for higher-than-standard cumulative clindamycin dose in Black versus White patient encounters. The pooled model included institution as a covariate, and heterogeneity across institutions was assessed with a race-by-site interaction test. Panel B shows the pooled adjusted OR after repeated analyses in which encounters with Unknown race/ethnicity were progressively reassigned toward the Black category. Larger values of the reassignment parameter indicate stronger reassignment toward the Black category. Panel C shows observed site-specific event rates among encounters with known versus Unknown race/ethnicity. Panel D shows site-specific unadjusted ORs comparing Unknown versus known race/ethnicity encounters. Higher-than-standard cumulative clindamycin dose was defined as >8,400 mg.

To evaluate the potential impact of excluding encounters with Unknown race/ethnicity, we performed sensitivity analyses in which progressively larger proportions of encounters with Unknown race/ethnicity were reassigned to the Black category and the primary model was re-estimated under each scenario. Although the estimated odds ratio attenuated with increasing reassignment, the direction of the association remained consistent across all scenarios (Figure 2B). The composition of encounters with Unknown race/ethnicity was heterogeneous and varied across institutions (Figure 2C–D; Figure S1).

Additional antibiotic exposure was documented elsewhere in the electronic health record among a subset of clindamycin-prescribing encounters. Of the 2,197 encounters involving dentist-prescribed clindamycin, 551 (25.1%) included documentation of at least one additional antibiotic exposure. Notably, 93.1% (513/551) of these encounters involved self-reported clindamycin, indicating that when concurrent antibiotic exposure was reported, it was almost always another clindamycin prescription (Table S4). The frequency of documented concurrent antibiotic exposure varied substantially across institutions, ranging from 3.4% at Site 1 to 31.9% at Site 4 after excluding Site 2, where no concurrent exposures were documented.

Finally, procedure mix among observations with higher-than-standard cumulative antibiotic dosing is shown in Figure S2. Although distributions differed across institutions and race/ethnicity groups, no consistent pattern suggested that measured procedure context alone explained the increased adjusted odds of higher-than-standard cumulative clindamycin dose observed among Black patient encounters.

## DISCUSSION

Overall antibiotic prescribing rates were low, consistent with other reports in the academic dental setting.^24^ Prior stewardship evaluations have nevertheless identified opportunities to improve antibiotic selection and duration.^24^ Higher-than-standard antibiotic exposure has also been documented in academic dental settings, and we observed a similar pattern in our cohort, where higher-than-standard cumulative amoxicillin exposure occurred in nearly one-third of amoxicillin encounters. We also identified higher-than-standard cumulative clindamycin exposure in a subset of clindamycin-prescribing encounters. Because clindamycin poses among the highest risks of CDI and poor outcomes, this finding has greater clinical significance than comparable exposure patterns involving lower-risk antibiotics.

Given well-documented racial/ethnic disparities in CDI incidence, severity, and outcomes, we determined whether cumulative exposure to a high-risk antibiotic differed across racial and ethnic groups. Black populations experience disproportionate burdens of severe CDI and adverse CDI outcomes. In our study, among encounters in which clindamycin was prescribed, Black patients were more than twice as likely to receive higher-than-standard cumulative clindamycin doses compared with White patients. Although our study did not evaluate CDI directly, this finding raises the possibility that differential exposure to a high-CDI-risk antibiotic may contribute to downstream disparities in CDI vulnerability and outcomes.

Regional prescribing practices and clinical context are well-established drivers of variation in antibiotic prescribing. Prior research has demonstrated substantial geographic variation in dental antibiotic prescribing, including clindamycin use, while racial and ethnic population composition also varies considerably across regions.^25–27^ Antibiotic use also differs across dental procedure types, and penicillin allergy is a recognized indication for selecting clindamycin when antibiotic therapy is needed.^11,24,28^ However, these factors do not necessarily explain differences in cumulative exposure once clindamycin is prescribed. In our study, sensitivity analyses did not suggest that the observed association between black patient encounters and higher-than-standard cumulative clindamycin was driven by a single institution or geographic region. Similarly, procedure distributions among encounters with higher-than-standard cumulative clindamycin exposure varied across institutions and racial and ethnic groups, but no consistent pattern emerged. Thus, other factors may have contributed to differences in cumulative clindamycin exposure.

Identifying factors associated with higher cumulative clindamycin exposure becomes even more important when considering that dentistry represents only one of several healthcare settings in which patients may be exposed to this antibiotic. Notably, when patients reported antibiotic exposures independent of the clindamycin-prescribing dental encounter, the reported antibiotic was almost always another clindamycin prescription. This pattern was observed across racial and ethnic groups and participating institutions, indicating that cumulative clindamycin exposure often extends beyond what is captured within dental prescribing records alone. However, antibiotic exposures occurring in different healthcare settings are not always documented within the same health record. Consistent with this challenge, documentation of patient-reported antibiotic exposure varied across participating institutions in our cohort. This highlights the difficulty of characterizing total clindamycin exposure when prescribing within a single clinical setting is evaluated in isolation, suggesting that dose-dependent CDI risk may be underestimated.

Similar exposure-related disparities have been reported for other CDI-associated medications, including proton pump inhibitors, where prolonged use and lower rates of medication de-escalation have been described among Black patients, suggesting that inequities in cumulative exposure may extend beyond antibiotic prescribing.^29–31^ However, clindamycin remains among the medications most strongly associated with CDI, making differences in cumulative exposure particularly relevant.

CDI remains a well-recognized adverse outcome of clindamycin exposure, but the consequences of clindamycin exposure also extend to other infectious outcomes. Prior clindamycin exposure has been implicated in increased susceptibility to, or severity of, a range of infections, including those involving *Salmonella* and *Klebsiella* species.^32,33^ Although these associations have been less extensively studied than CDI, Black and Hispanic populations have repeatedly been shown to experience more severe infection and worse clinical outcomes across several of these conditions.^34–38^ These broader downstream consequences further underscore the importance of understanding patterns of cumulative exposure to high-risk antibiotics.

This study has several limitations. Participating institutions were academic clinics, and our findings may not be generalizable to other dental care settings. Our analysis also relied on electronic health record data containing missing documentation of race and ethnicity. Because data were likely not missing at random and documentation practices varied across institutions, some degree of bias may persist despite extensive sensitivity analyses.^39,40^ In addition, electronic health records capture prescribing practices but do not offer any insight into medication adherence, which prevents us from conducting a direct assessment of true antibiotic exposure. Finally, several analytic simplifications were necessary to harmonize data across institutions: some antibiotics were grouped into broader categories, potentially obscuring differences between the individual agents; self-reported antibiotic exposure could not be incorporated into primary adjusted models; and residual confounding from unmeasured clinical, social, or structural factors may be present.

## CONCLUSION

Despite these limitations, this study offers actionable insights for antimicrobial stewardship efforts. Although overall antibiotic prescribing rates were relatively low, as expected in academic dental settings where stewardship practices are commonly emphasized, disparities in exposure to a high-risk antibiotic emerged when cumulative exposure was evaluated. As such, stewardship metrics focused solely on antibiotic receipt or antibiotic selection may overlook clinically meaningful differences in exposure that could contribute to disparate vulnerability. Incorporating measures of cumulative exposure may help identify prescribing inequities that are not apparent when antibiotic use is evaluated primarily as a binary outcome and may provide additional opportunities for targeted stewardship interventions. More broadly, because racial and ethnic disparities in infection susceptibility and disease severity are well documented, identifying disparities in modifiable exposure-related infection risk factors may help inform stewardship strategies that advance both antibiotic safety and health equity.

## Data Availability

All data produced in the present study are available upon reasonable request to the authors

## Acknowledgements

We acknowledge the BigMouth Dental Data Repository.

---

**Figure S1.**
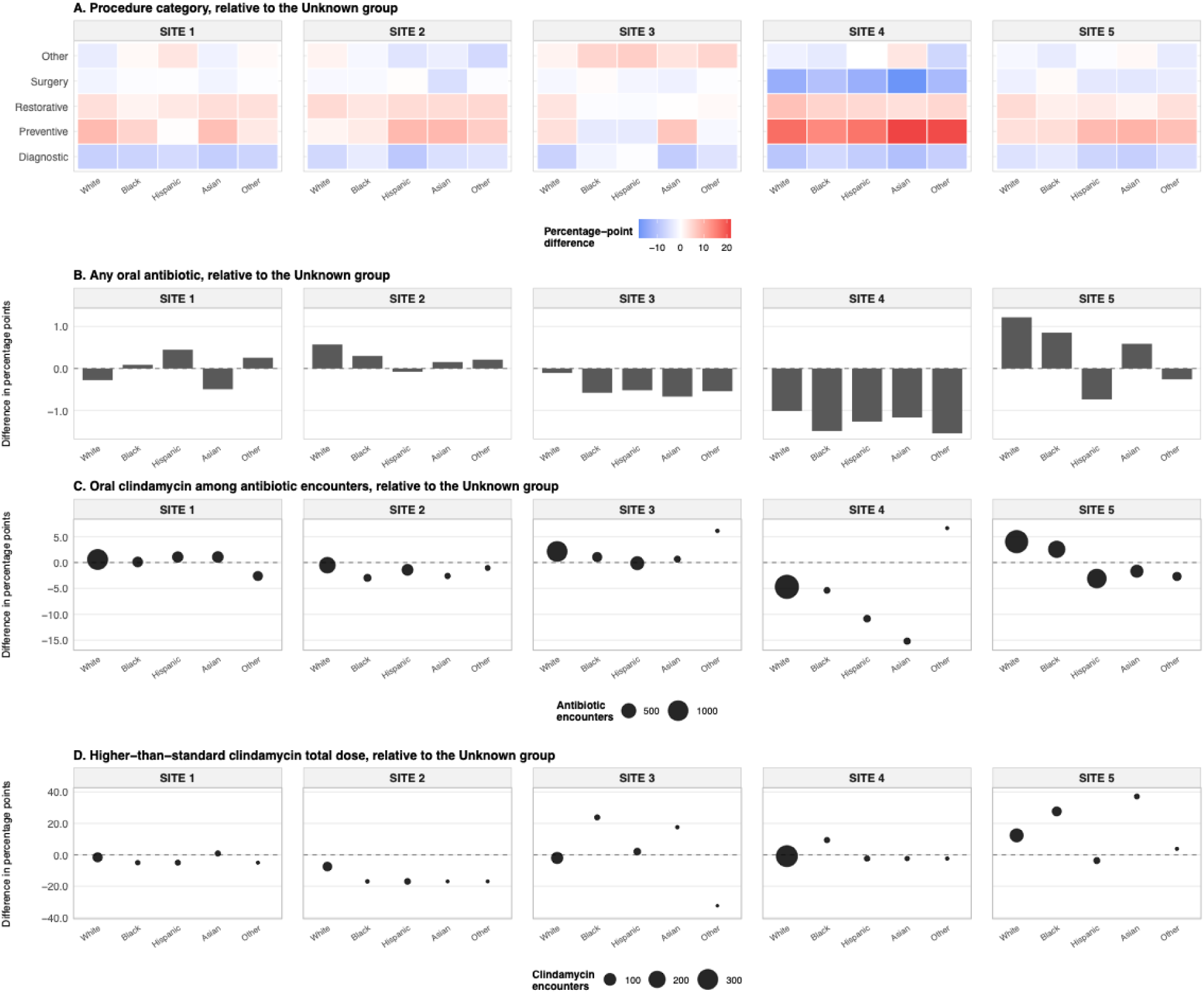
Encounter-level differences relative to Unknown race/ethnicity within institution. Panel A shows differences in the distribution of primary procedure categories relative to the Unknown race/ethnicity group within each institution. Panel B shows percentage-point differences in the proportion of patient encounters with any oral antibiotic. Panel C shows percentage-point differences in the proportion of encounters with an oral antibiotic prescription where clindamycin was selected. Panel D shows percentage-point differences in the proportion of clindamycin prescribed by dental providers with higher-than-standard total dose. Positive values indicate higher values than the Unknown race/ethnicity group within the same institution. Point sizes in Panels C and D are proportional. Higher-than-standard clindamycin dosing was defined as total oral clindamycin dose >8,400 mg.

**Figure S2.**
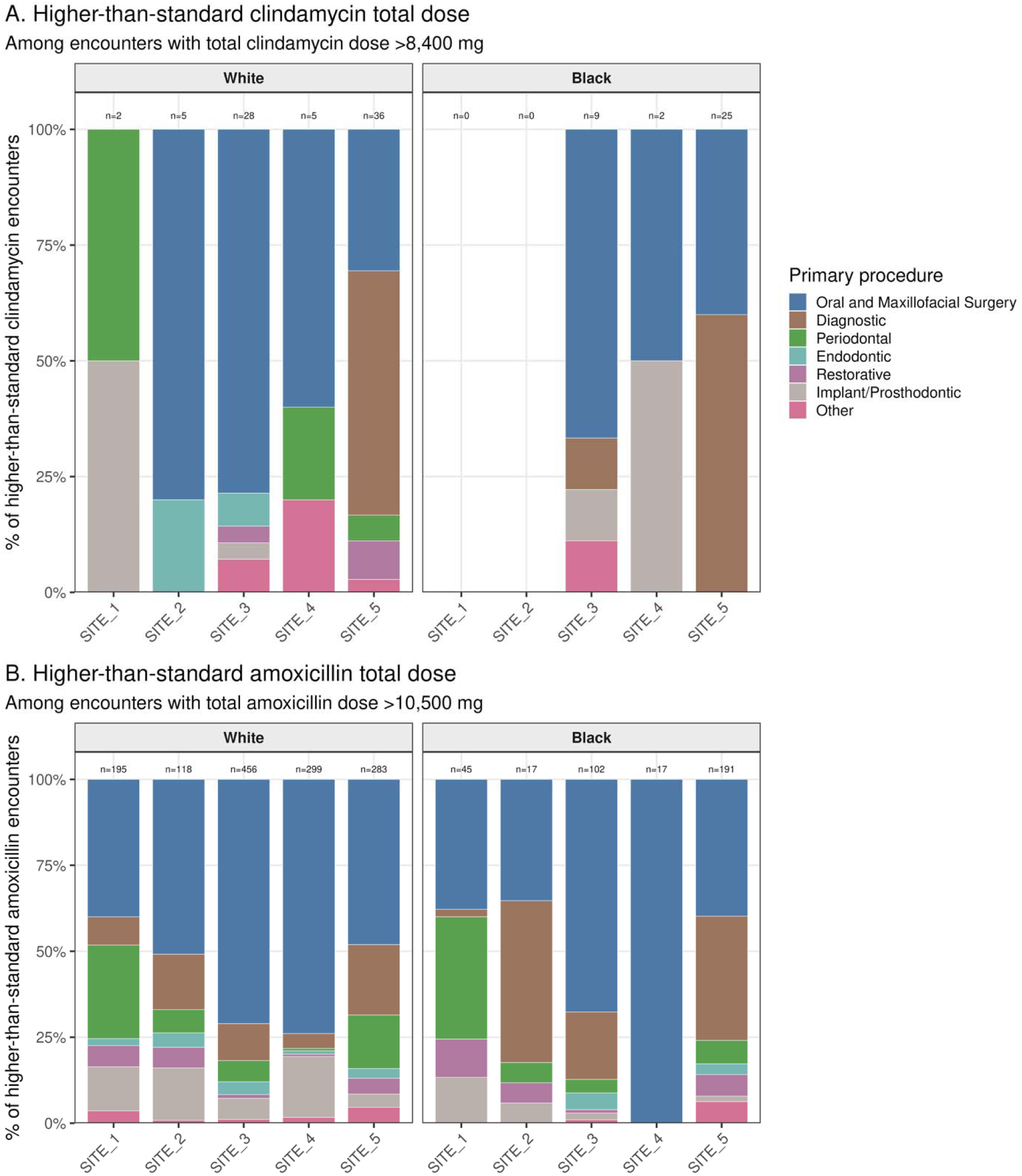
Primary procedure mix among higher-than-standard antibiotic encounters by site and race/ethnicity. Panel A shows the distribution of primary procedure categories among higher-than-standard clindamycin encounters (total clindamycin dose >8,400 mg), and Panel B shows the corresponding distribution among higher-than-standard amoxicillin encounters (total amoxicillin dose >10,500 mg). Bars are stratified by institution and race/ethnicity and display the percentage of higher-than-standard encounters attributable to each primary procedure category within each site-by-race/ethnicity stratum. Numbers above bars indicate the number of higher-than-standard encounters in each stratum. Findings should be interpreted descriptively because several site-by-race/ethnicity strata included small numbers of higher-than-standard clindamycin encounters.

**Table S1.**
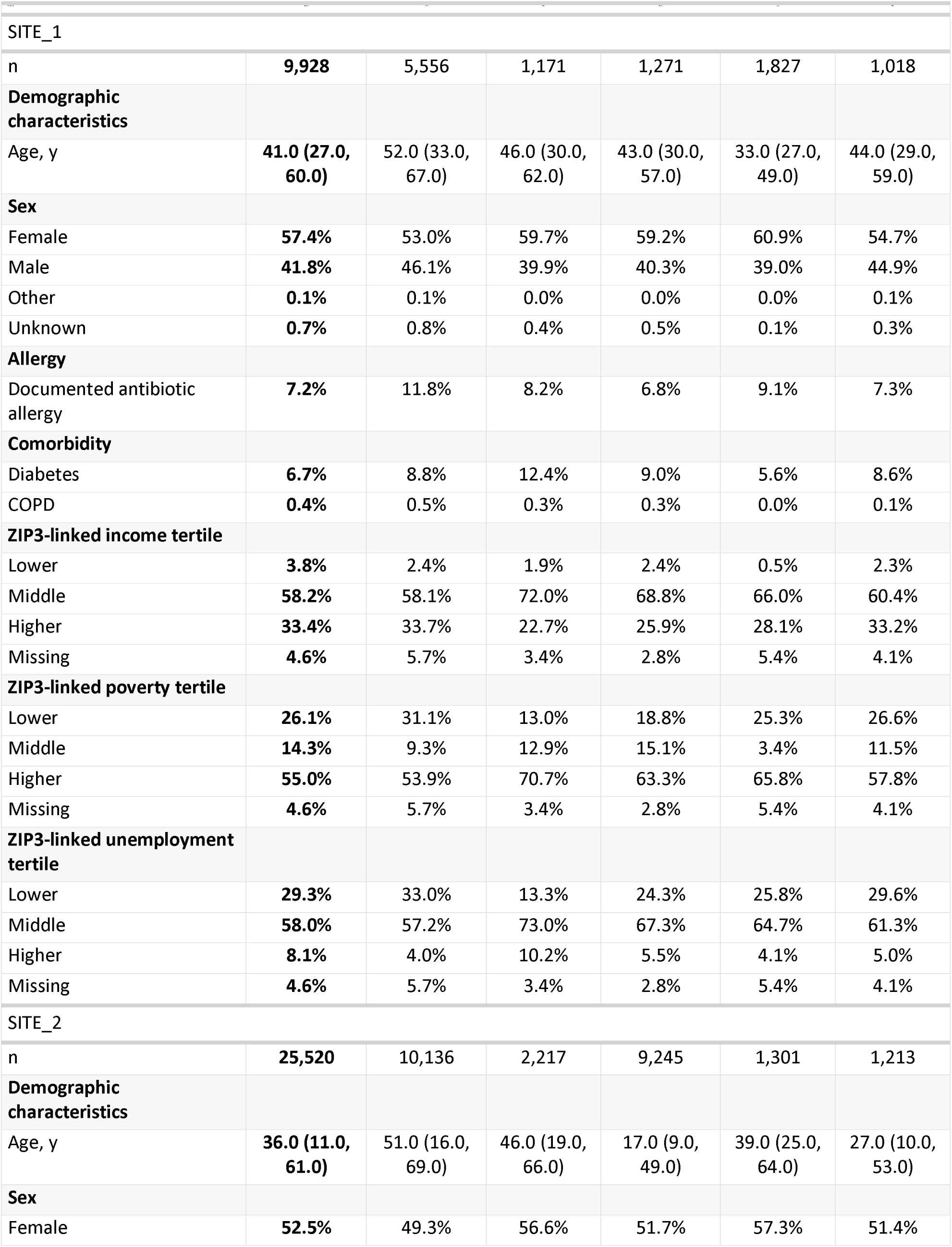
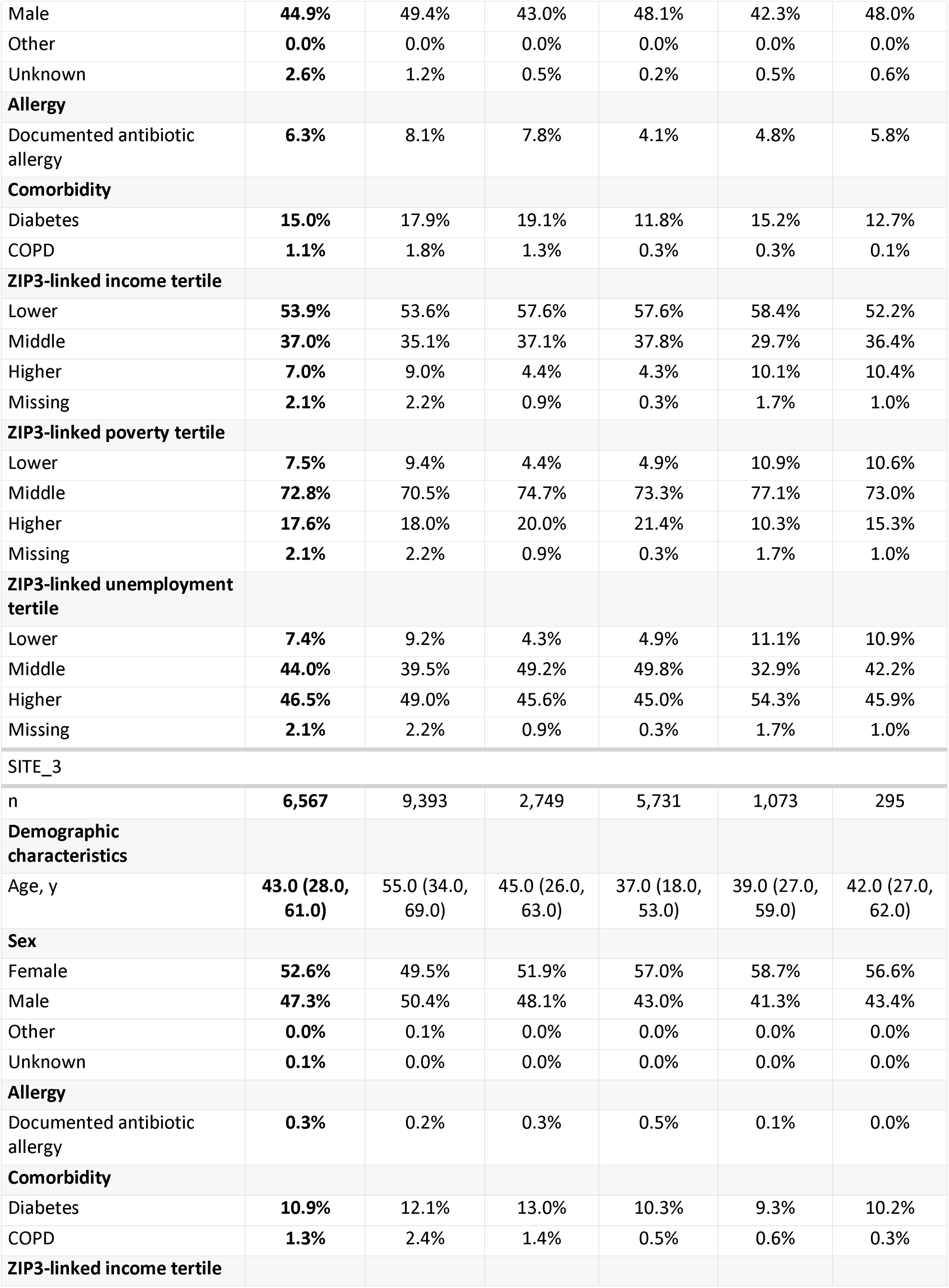
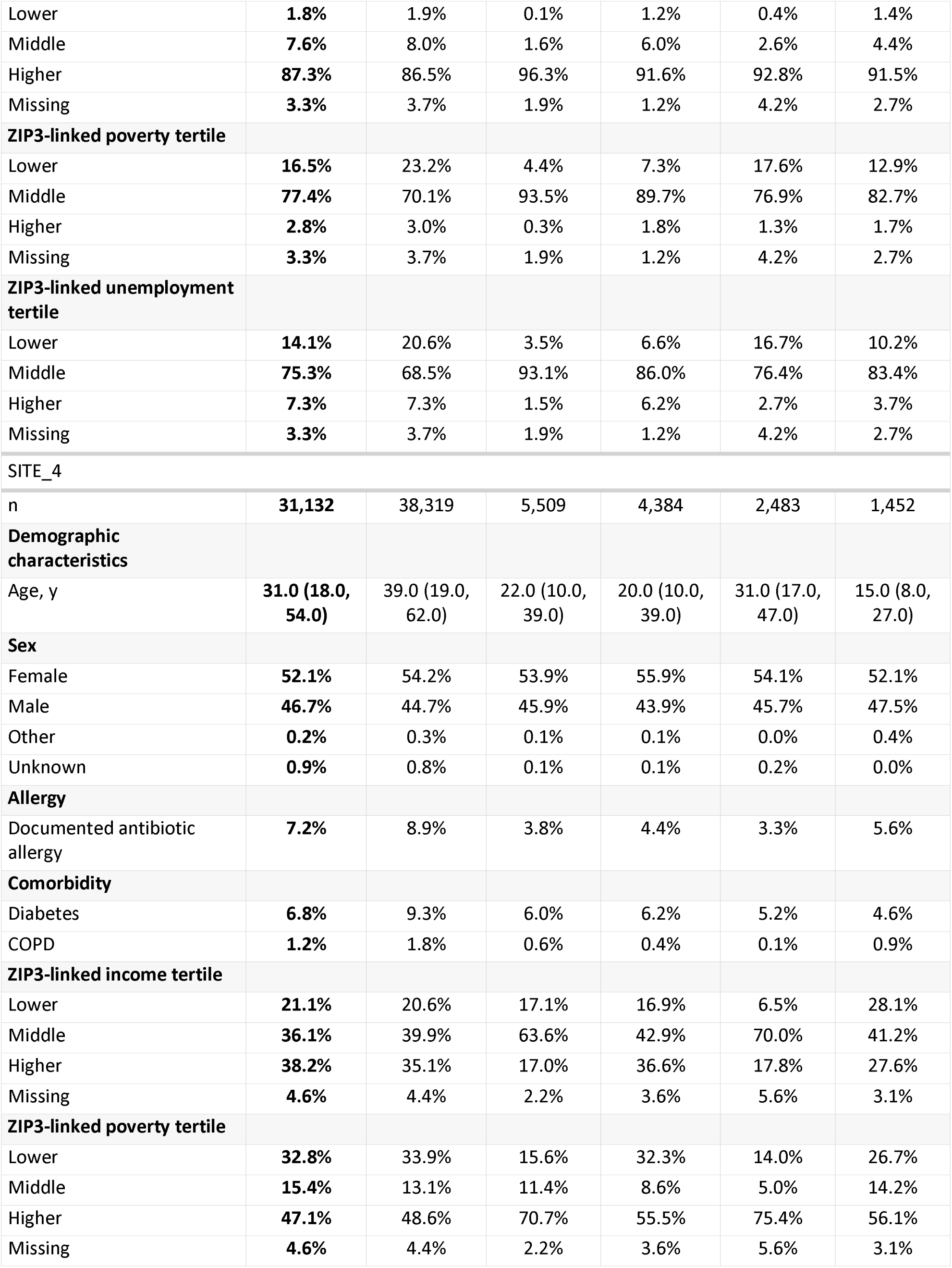
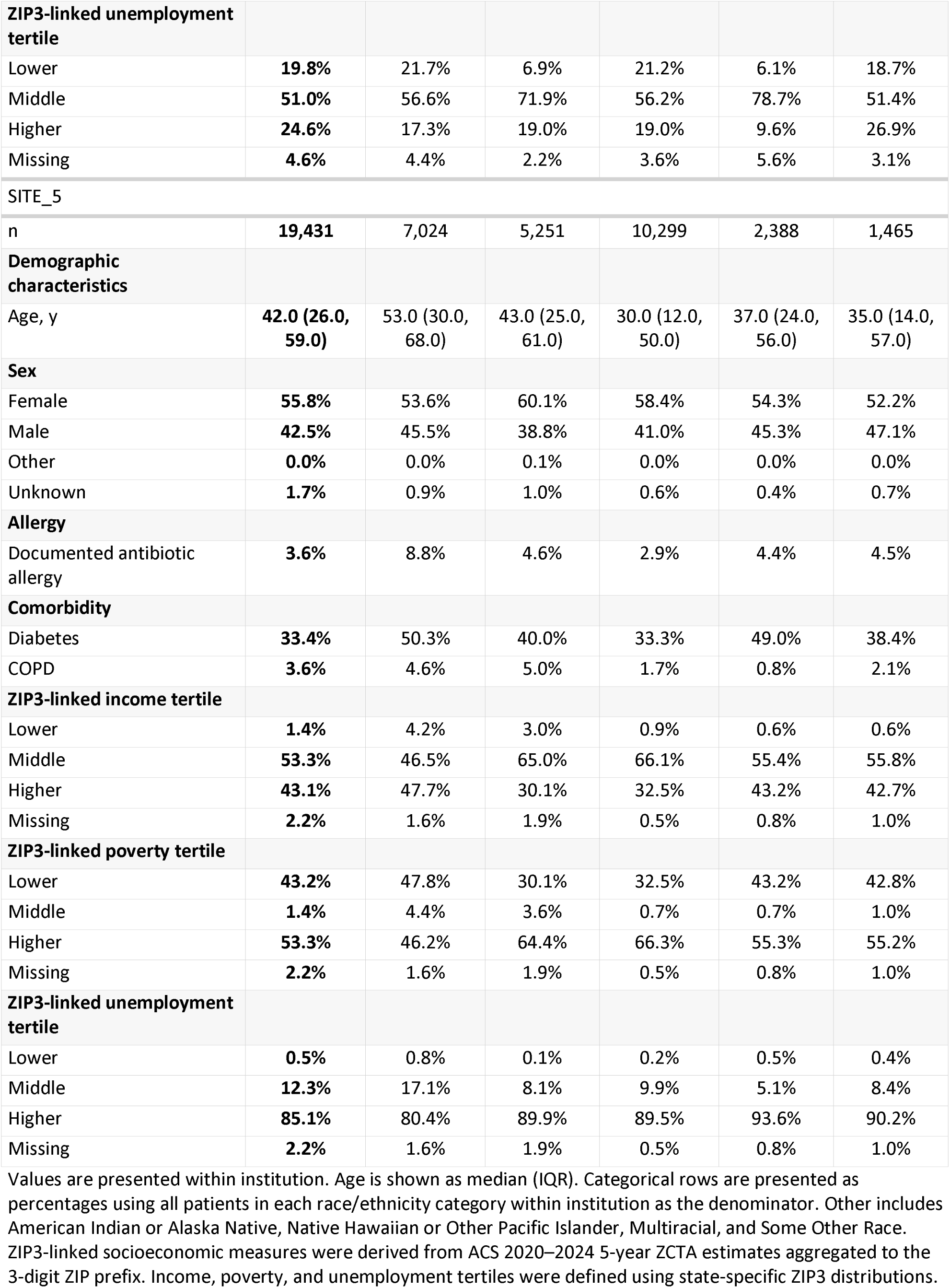
Patient-level characteristics by race/ethnicity within institution.

**Table S2.**
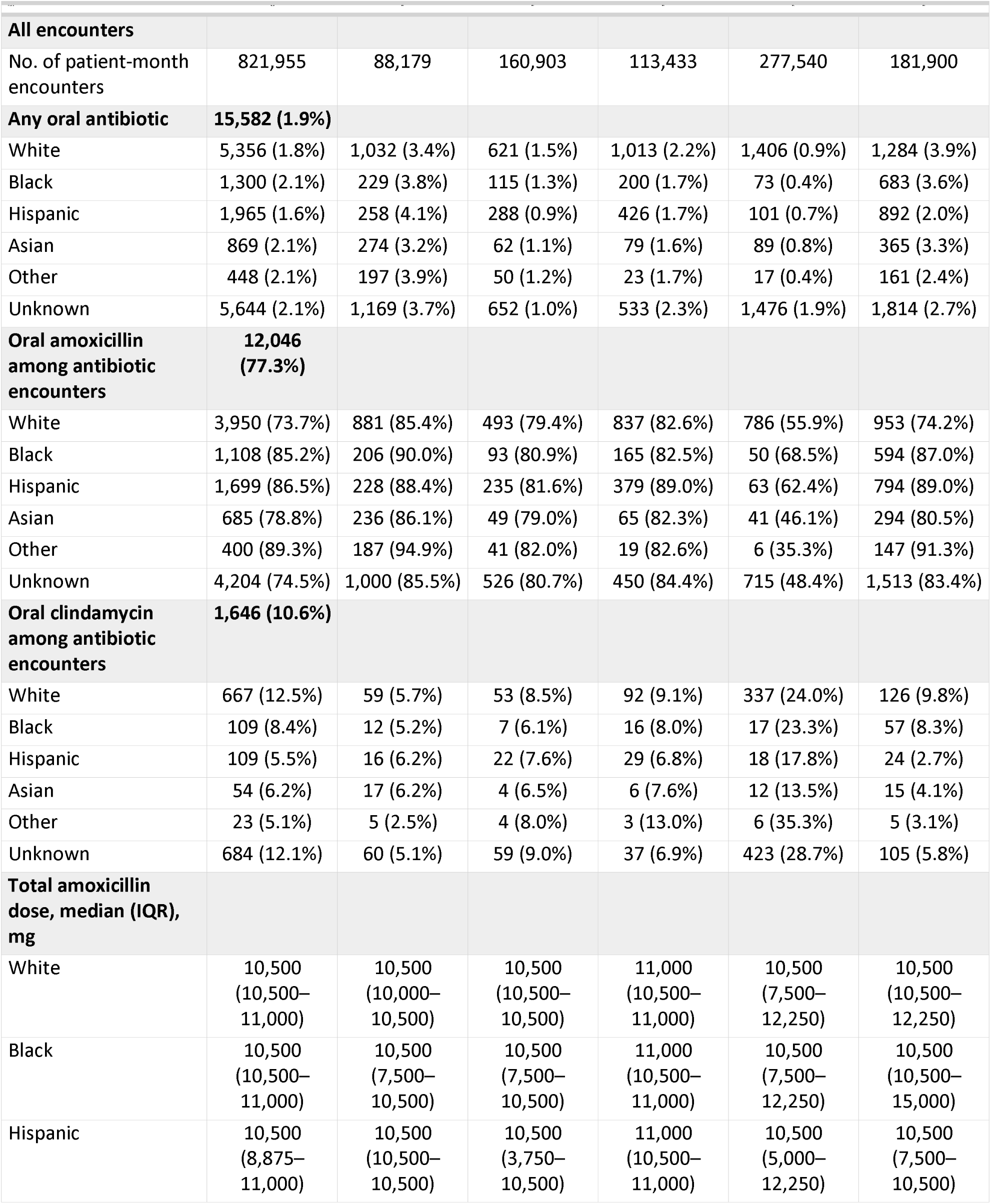
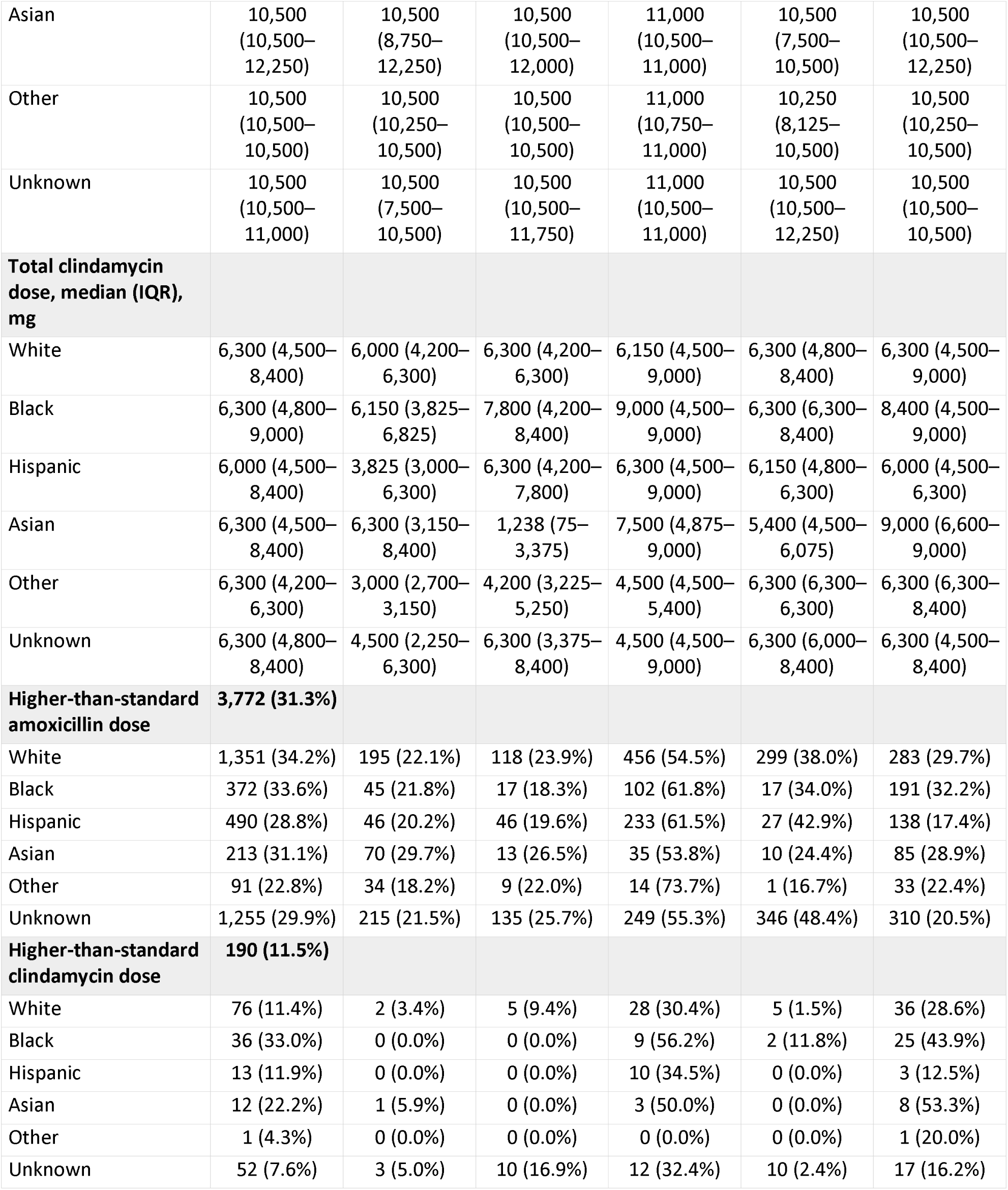
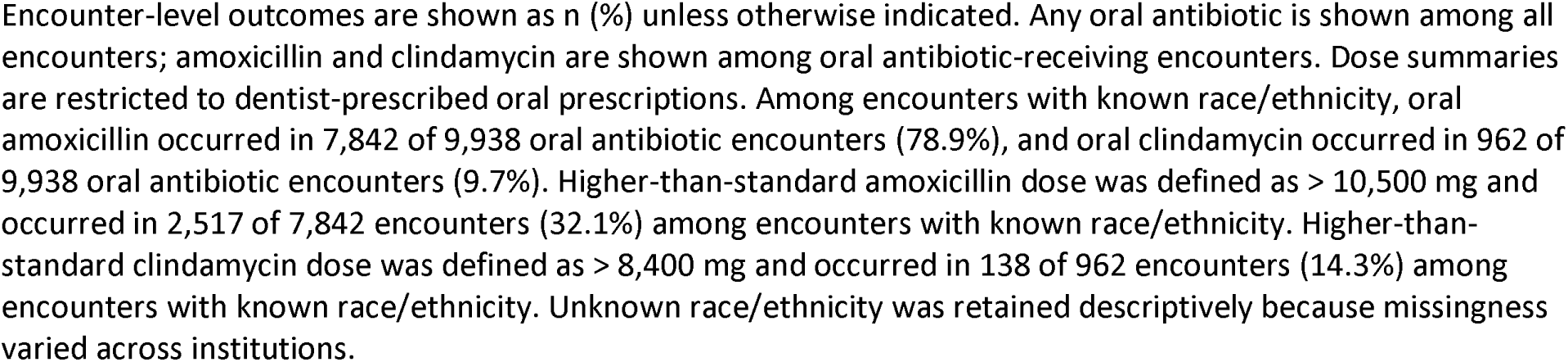
Encounter-level descriptive antibiotic prescribing outcomes by race/ethnicity and institution.

**Table S3.** Full multivariable model results for encounter-level antibiotic prescribing outcomes.

| Outcome | Model N | Variable | Adjusted OR (95% CI) | P value |
| --- | --- | --- | --- | --- |
| **Any oral antibiotic** | **552,428** |  |  |  |
|  |  | Age, per year | 1.01 (1.01–1.01) | <0.001 |
|  |  | Calendar year: 2022 vs 2021 | 0.87 (0.83–0.91) | <0.001 |
|  |  | Calendar year: 2023 vs 2021 | 0.83 (0.79–0.87) | <0.001 |
|  |  | Institution: SITE 2 vs SITE 1 | 0.23 (0.21–0.25) | <0.001 |
|  |  | Institution: SITE 3 vs SITE 1 | 0.48 (0.45–0.52) | <0.001 |
|  |  | Institution: SITE 4 vs SITE 1 | 0.18 (0.17–0.19) | <0.001 |
|  |  | Institution: SITE 5 vs SITE 1 | 0.73 (0.69–0.78) | <0.001 |
|  |  | Primary procedure category: Diagnostic | 2.05 (1.70–2.50) | <0.001 |
|  |  | Primary procedure category: Endodontic | 4.25 (3.45–5.29) | <0.001 |
|  |  | Primary procedure category: Implant/Prosthodontic | 12.41 (10.21–15.21) | <0.001 |
|  |  | Primary procedure category: Oral and Maxillofacial Surgery | 23.63 (19.71–28.65) | <0.001 |
|  |  | Primary procedure category: Orthodontics | 0.04 (0.01–0.08) | <0.001 |
|  |  | Primary procedure category: Other/Unknown | 0.00 (0.00–0.00) | 0.871 |
|  |  | Primary procedure category: Periodontal | 3.72 (3.09–4.53) | <0.001 |
|  |  | Primary procedure category: Preventive | 0.62 (0.50–0.77) | <0.001 |
|  |  | Primary procedure category: Prosthodontic | 0.90 (0.67–1.20) | 0.486 |
|  |  | Primary procedure category: Restorative | 0.90 (0.73–1.12) | 0.346 |
|  |  | Race/ethnicity: Asian vs White | 1.03 (0.95–1.11) | 0.444 |
|  |  | Race/ethnicity: Black vs White | 0.85 (0.79–0.91) | <0.001 |
|  |  | Race/ethnicity: Hispanic vs White | 0.80 (0.75–0.84) | <0.001 |
|  |  | Race/ethnicity: Other vs White | 0.87 (0.78–0.96) | 0.009 |
|  |  | Sex: Male | 0.95 (0.91–0.99) | 0.021 |
|  |  | Sex: NULL | 0.00 (NA–1151.76) | 0.971 |
|  |  | Sex: Others | 0.55 (0.17–1.30) | 0.236 |
|  |  | Sex: Unknown | 0.14 (0.02–0.45) | 0.007 |
| **Oral clindamycin among antibiotic encounters** | **9,895** |  |  |  |
|  |  | Age, per year | 1.00 (0.99–1.00) | 0.384 |
|  |  | Calendar year: 2022 vs 2021 | 0.88 (0.74–1.04) | 0.124 |
|  |  | Calendar year: 2023 vs 2021 | 0.67 (0.56–0.80) | <0.001 |
|  |  | Institution: SITE 2 vs SITE 1 | 1.20 (0.88–1.62) | 0.245 |
|  |  | Institution: SITE 3 vs SITE 1 | 1.49 (1.14–1.95) | 0.003 |
|  |  | Institution: SITE 4 vs SITE 1 | 4.35 (3.44–5.55) | <0.001 |
|  |  | Institution: SITE 5 vs SITE 1 | 1.21 (0.96–1.55) | 0.117 |
|  |  | Primary procedure category: Diagnostic | 0.71 (0.40–1.32) | 0.250 |
|  |  | Primary procedure category: Endodontic | 0.52 (0.26–1.06) | 0.063 |
|  |  | Primary procedure category: Implant/Prosthodontic | 0.60 (0.34–1.13) | 0.094 |
|  |  | Primary procedure category: Oral and Maxillofacial Surgery | 0.56 (0.33–1.03) | 0.047 |
|  |  | Primary procedure category: Orthodontics | 0.00 (NA–3055.54) | 0.955 |
|  |  | Primary procedure category: Periodontal | 0.38 (0.21–0.72) | 0.002 |
|  |  | Primary procedure category: Preventive | 0.33 (0.16–0.67) | 0.002 |
|  |  | Primary procedure category: Prosthodontic | 0.41 (0.14–1.11) | 0.092 |
|  |  | Primary procedure category: Restorative | 0.78 (0.42–1.53) | 0.456 |
|  |  | Race/ethnicity: Asian vs White | 0.60 (0.44–0.80) | <0.001 |
|  |  | Race/ethnicity: Black vs White | 0.85 (0.67–1.06) | 0.163 |
|  |  | Race/ethnicity: Hispanic vs White | 0.55 (0.43–0.68) | <0.001 |
|  |  | Race/ethnicity: Other vs White | 0.57 (0.36–0.86) | 0.012 |
|  |  | Sex: Male | 0.57 (0.49–0.66) | <0.001 |
|  |  | Sex: Others | 0.00 (NA–46149038.94) | 0.961 |
|  |  | Sex: Unknown | 0.00 (NA–540274561315755261952.00) | 0.976 |
| **Higher-than-standard amoxicillin total dose** | **7,807** |  |  |  |
|  |  | Age, per year | 1.00 (0.99–1.00) | 0.007 |
|  |  | Calendar year: 2022 vs 2021 | 1.68 (1.48–1.91) | <0.001 |
|  |  | Calendar year: 2023 vs 2021 | 1.86 (1.64–2.11) | <0.001 |
|  |  | Institution: SITE 2 vs SITE 1 | 1.22 (0.99–1.50) | 0.055 |
|  |  | Institution: SITE 3 vs SITE 1 | 4.10 (3.49–4.82) | <0.001 |
|  |  | Institution: SITE 4 vs SITE 1 | 1.88 (1.56–2.26) | <0.001 |
|  |  | Institution: SITE 5 vs SITE 1 | 1.13 (0.98–1.32) | 0.093 |
|  |  | Primary procedure category: Diagnostic | 2.17 (1.30–3.77) | 0.004 |
|  |  | Primary procedure category: Endodontic | 1.13 (0.64–2.07) | 0.671 |
|  |  | Primary procedure category: Implant/Prosthodontic | 1.38 (0.81–2.41) | 0.249 |
|  |  | Primary procedure category: Oral and Maxillofacial Surgery | 1.56 (0.95–2.66) | 0.091 |
|  |  | Primary procedure category: Orthodontics | 1.71 (0.22–9.75) | 0.561 |
|  |  | Primary procedure category: Periodontal | 1.07 (0.64–1.86) | 0.791 |
|  |  | Primary procedure category: Preventive | 0.78 (0.43–1.45) | 0.429 |
|  |  | Primary procedure category: Prosthodontic | 1.23 (0.52–2.86) | 0.636 |
|  |  | Primary procedure category: Restorative | 1.36 (0.77–2.45) | 0.298 |
|  |  | Race/ethnicity: Asian vs White | 1.06 (0.88–1.28) | 0.541 |
|  |  | Race/ethnicity: Black vs White | 1.06 (0.90–1.23) | 0.494 |
|  |  | Race/ethnicity: Hispanic vs White | 0.74 (0.64–0.85) | <0.001 |
|  |  | Race/ethnicity: Other vs White | 0.80 (0.62–1.03) | 0.084 |
|  |  | Sex: Male | 0.99 (0.89–1.09) | 0.809 |
|  |  | Sex: Others | 0.75 (0.03–8.27) | 0.816 |
|  |  | Sex: Unknown | 0.00 (NA–108430.20) | 0.942 |
| **Higher-than-standard clindamycin total dose** | **959** |  |  |  |
|  |  | Age, per year | 0.99 (0.98–1.01) | 0.269 |
|  |  | Calendar year: 2022 vs 2021 | 2.67 (1.56–4.66) | <0.001 |
|  |  | Calendar year: 2023 vs 2021 | 3.20 (1.78–5.85) | <0.001 |
|  |  | Institution: SITE 2 vs SITE 1 | 4.02 (0.90–21.11) | 0.073 |
|  |  | Institution: SITE 3 vs SITE 1 | 16.36 (5.39–71.72) | <0.001 |
|  |  | Institution: SITE 4 vs SITE 1 | 0.60 (0.15–2.92) | 0.476 |
|  |  | Institution: SITE 5 vs SITE 1 | 15.30 (5.26–65.33) | <0.001 |
|  |  | Primary procedure category: Diagnostic | 1.55 (0.43–6.48) | 0.521 |
|  |  | Primary procedure category: Endodontic | 0.74 (0.13–4.19) | 0.726 |
|  |  | Primary procedure category: Implant/Prosthodontic | 0.97 (0.20–4.95) | 0.969 |
|  |  | Primary procedure category: Oral and Maxillofacial Surgery | 0.83 (0.24–3.35) | 0.773 |
|  |  | Primary procedure category: Periodontal | 0.59 (0.12–2.97) | 0.507 |
|  |  | Primary procedure category: Preventive | 0.17 (0.01–1.54) | 0.154 |
|  |  | Primary procedure category: Prosthodontic | 0.91 (0.03–13.13) | 0.949 |
|  |  | Primary procedure category: Restorative | 0.70 (0.14–3.70) | 0.664 |
|  |  | Race/ethnicity: Asian vs White | 2.15 (0.90–5.00) | 0.077 |
|  |  | Race/ethnicity: Black vs White | 2.19 (1.25–3.82) | 0.006 |
|  |  | Race/ethnicity: Hispanic vs White | 0.58 (0.28–1.14) | 0.126 |
|  |  | Race/ethnicity: Other vs White | 0.30 (0.02–1.65) | 0.260 |
|  |  | Sex: Male | 0.72 (0.45–1.14) | 0.165 |

White patient encounters were the reference group for race/ethnicity. Reference groups were SITE 1 for institution and 2021 for calendar year. Models were adjusted for age, sex, institution, calendar year, and primary procedure category. The oral clindamycin model was restricted to oral antibiotic-receiving encounters. Higher-than-standard amoxicillin total dose was defined as total oral amoxicillin dose > 10,500 mg. Higher-than-standard clindamycin total dose was defined as total oral clindamycin dose > 8,400 mg. Self-reported antibiotic entries were excluded from dose-based analyses. Analyses were restricted to encounters with known race/ethnicity.

**Table S4.** Self-reported antibiotic co-exposure among dentist-prescribed oral clindamycin encounters.

|  |  |  |  |
| --- | --- | --- | --- |
| **Overall** | **2,197** | **551 (25.1%)** | **513 (23.4%)** |
| **Race/ethnicity** |  |  |  |
| White | 936 | 269 (28.7%) | 256 (27.4%) |
| Black | 149 | 40 (26.8%) | 38 (25.5%) |
| Hispanic | 146 | 37 (25.3%) | 30 (20.5%) |
| Asian | 65 | 11 (16.9%) | 11 (16.9%) |
| Other | 34 | 11 (32.4%) | 10 (29.4%) |
| Unknown | 867 | 183 (21.1%) | 168 (19.4%) |
| **Institution** |  |  |  |
| SITE_1 | 175 | 6 (3.4%) | 4 (2.3%) |
| SITE_2 | 149 | 0 (0.0%) | 0 (0.0%) |
| SITE_3 | 262 | 79 (30.2%) | 76 (29.0%) |
| SITE_4 | 1,193 | 380 (31.9%) | 351 (29.4%) |
| SITE_5 | 418 | 86 (20.6%) | 82 (19.6%) |

Values are shown as n (%) unless otherwise indicated. Denominator for each row is the number of encounter-level observations with dentist-prescribed oral clindamycin. Self-reported antibiotic variables reflect concurrent documentation of patient-reported antibiotic exposure within the same patient-month encounter. Race/ethnicity categories were harmonized for analysis.

